# Real-world evaluation of deep learning algorithms to classify functional pathogenic germline variants

**DOI:** 10.1101/2024.04.05.24305402

**Authors:** Ryan D. Chow, Ravi B. Parikh, Katherine L. Nathanson

**Affiliations:** Department of Medicine, Hospital of the University of Pennsylvania, Philadelphia, PA, USA; Division of Health Policy, Perelman School of Medicine, University of Pennsylvania, Philadelphia, PA, USA; Penn Center for Cancer Care Innovation, Abramson Cancer Center, Philadelphia, PA, USA; Division of Hematology and Oncology, Perelman School of Medicine, University of Pennsylvania, Philadelphia, PA, USA; Corporal Michael J. Crescenz VA Medical Center, Philadelphia, PA, USA; Basser Center for BRCA, Abramson Cancer Center, Perelman School of Medicine, University of Pennsylvania, Philadelphia, PA, USA; Division of Translational Medicine and Human Genetics, Department of Medicine, Perelman School of Medicine, University of Pennsylvania, Philadelphia, PA, USA

## Abstract

Deep learning models for variant pathogenicity prediction can recapitulate expert-curated annotations, but their performance remains unexplored on actual disease phenotypes in a real-world setting. Here, we apply three state-of-the-art pathogenicity prediction models to classify hereditary breast cancer gene variants in the UK Biobank. Predicted pathogenic variants in *BRCA1, BRCA2* and *PALB2*, but not *ATM* and *CHEK2*, were associated with increased breast cancer risk. We explored gene-specific score thresholds for variant pathogenicity, finding that they could improve model performance. However, when specifically tasked with classifying variants of uncertain significance, the deep learning models were generally of limited clinical utility.

## Main Text

A core challenge of germline genetic testing is to distinguish variants that are pathogenic from those that are benign^1^. In the case of hereditary breast cancer genes such as *BRCA1* and *BRCA2*, this decision point has significant clinical ramifications, as patients carrying pathogenic variants may benefit from more aggressive screening and early intervention^2,3^. While extensive efforts have been devoted to cataloging and classifying variants in *BRCA1* and *BRCA2*, over 5% of identified alterations in these genes are nevertheless reported as variants of uncertain significance (VUSs).^4^ This issue is further amplified for hereditary breast cancer genes that have been comparatively less well-characterized, such as *ATM, CHEK2*, and *PALB2*. As current guidelines recommend against using VUSs to guide medical management^5,6^, ascertaining the pathogenicity of VUSs is a critical bottleneck limiting the utility of germline genetic testing.

Deep learning models for variant pathogenicity prediction have recently been developed that can independently recapitulate known pathogenicity annotations from gold-standard references such as the expert-curated ClinVar database^7–9^. The value proposition of these models is their potential to predict the pathogenicity of variants that have not yet been clinically studied or otherwise annotated in existing databases, thus potentially offering a scalable approach to close the interpretation gap for VUSs. However, model-based pathogenicity predictions have yet to be evaluated in relation to the clinically relevant outcomes that would ultimately matter to patients – for instance, whether VUSs predicted to be pathogenic are functionally associated with increased risk of disease.

Here, we applied three state-of-the-art deep learning models for pathogenicity prediction (AlphaMissense^7^, EVE^8^, and ESM1b^9^) to classify germline missense variants in participants of the UK Biobank. The UK Biobank is a cohort of approximately 500,000 individuals with matched genomic profiling and longitudinal health record data, including cancer diagnosis history^10,11^. We benchmarked model-based pathogenicity predictions for five hereditary breast cancer genes^12^ (*BRCA1, BRCA2, ATM, CHEK2*, and *PALB2*) against ClinVar annotations, evaluating whether current deep learning models are able to distinguish pathogenic germline variants that functionally confer increased risk of breast cancer in a clinically relevant context.

We identified 469,623 UK Biobank participants with matched exome sequencing data and health record data. We compiled all unique missense variants in *BRCA1, BRCA2, ATM, CHEK2*, and *PALB2* among UK Biobank participants (**Table S1**). Classifying each missense variant based on ClinVar annotations, we observed that the majority were VUSs – ranging from 63.7% of *BRCA1* variants to 96.7% of *CHEK2* variants (**Figure 1a**). Consolidating the variant calls to the participant-level, we observed that most participants with missense variants in a hereditary breast cancer gene were classified as benign variant carriers (**Figure 1b, Table S1**). The notable exception was *CHEK2*, as 88.2% of participants with *CHEK2* missense variants were labeled as VUS carriers. The divergence between the variant-level and participant-level analyses reflects the wide variation in allele frequencies across missense variants – i.e., benign variants in hereditary breast cancer genes are comparatively overrepresented in the UK Biobank population compared to pathogenic variants or VUSs.

**Figure 1:**
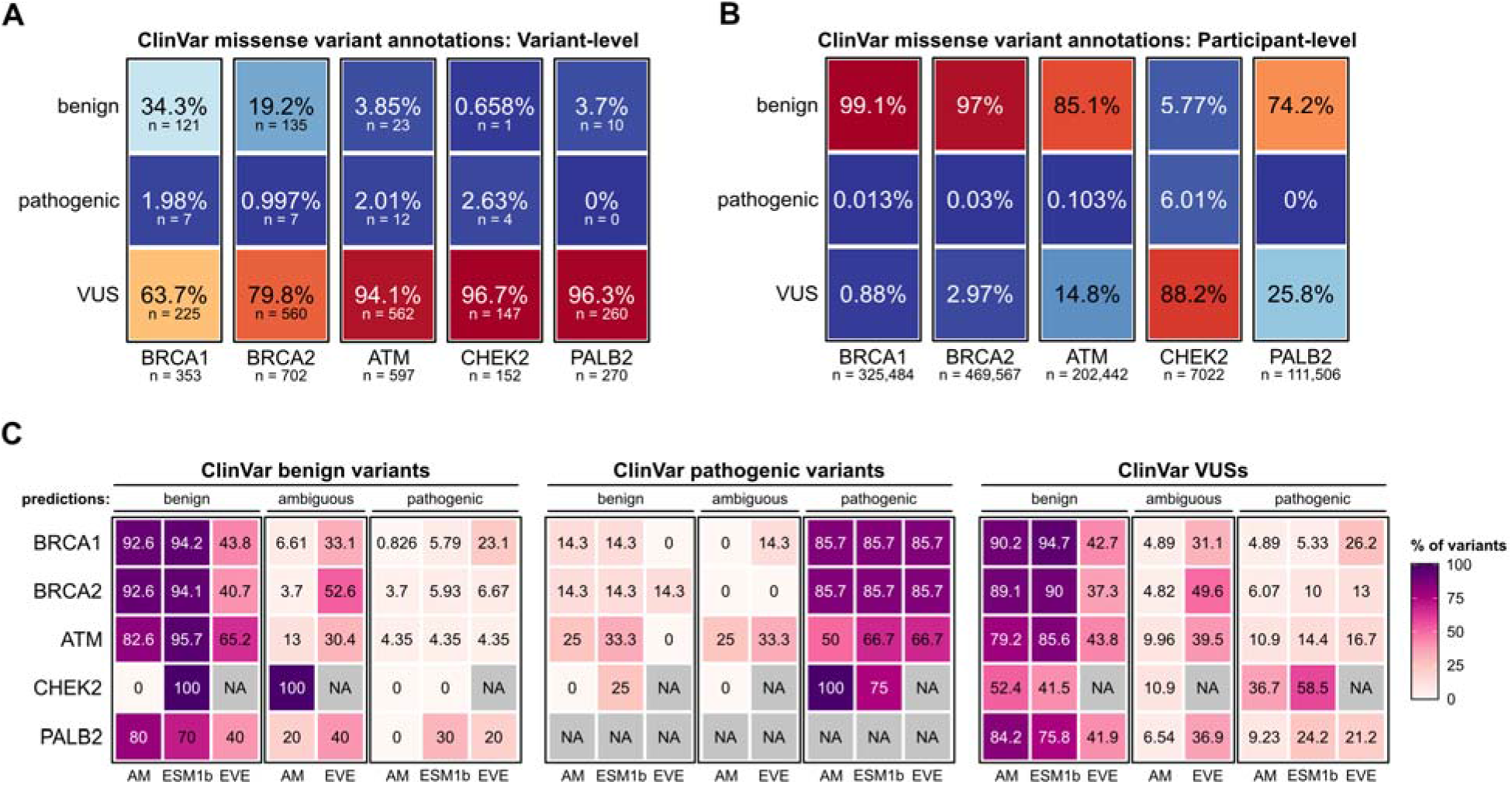
Deep learning models recapitulate ClinVar variant pathogenicity annotations for hereditary breast cancer genes. **A**. ClinVar classification of *BRCA1, BRCA2, ATM, CHEK2*, and *PALB2* missense variants in the UK Biobank cohort. Data are expressed as the percentage of total variants for each gene. The number of unique variants in each category is shown below. **B**. Participant-level classification of *BRCA1, BRCA2, ATM, CHEK2*, and *PALB2* missense variants carriers in the UK Biobank cohort. Data are expressed as the percentage of total variant carriers for each gene, with the total number of variant carriers for each gene shown below. **C**. Comparison of model-based pathogenicity predictions in reference to ClinVar annotations. Data are expressed as percentages for each gene, stratified by ClinVar annotation category (benign, pathogenic, or VUS). Thus, the percentages for each classifier sum to 100% within each ClinVar annotation category. AM, AlphaMissense.

We next compared the ability of each deep learning model to recapitulate benign ClinVar variant annotations (**Table S2**). AlphaMissense and ESM1b correctly labeled the majority of ClinVar benign variants, consistently outperforming EVE across the five hereditary breast cancer genes (**Figure 1c**). For instance, AlphaMissense correctly classified 92.6% of benign *BRCA1* missense variants, compared to 94.2% for ESM1b and 43.8% for EVE. However, model performance varied across genes -- AlphaMissense correctly labeled 80% of benign *PALB2* variants, compared to 70% for ESM1b and 40% for EVE. For the single benign *CHEK2* variant, AlphaMissense incorrectly annotated it as an ambiguous variant, while ESM1b correctly annotated it as a benign variant; *CHEK2* variant predictions were not included in the EVE database and thus were excluded from analysis. We similarly observed that the deep learning models were generally able to recapitulate pathogenic ClinVar variant annotations (**Figure 1c**). As an example, AlphaMissense, ESM1b, and EVE all correctly annotated 85.7% of ClinVar pathogenic *BRCA1* and *BRCA2* variants. However, we again noted variation in model performance across genes, as AlphaMissense correctly labeled only 50% of pathogenic *ATM* variants, compared to 66.7% for ESM1b and EVE.

Given that deep learning models would be most clinically useful for the classification of VUSs, we further assessed pathogenicity predictions from each of the models when applied to ClinVar VUSs. Overall, the majority of ClinVar VUSs were predicted to be benign variants by AlphaMissense and ESM1b, while EVE classifications were evenly split between benign and ambiguous designations. For instance, AlphaMissense classified 90.2% of *BRCA1* missense VUSs as benign, compared to 94.7% for ESM1b and 42.7% for EVE.

While prior studies on variant pathogenicity prediction have largely relied on ClinVar variant annotations as the gold-standard benchmark for model evaluation, we sought to leverage the linked clinicogenomic data in the UK Biobank to conduct a real-world assessment of whether the deep learning models could identify pathogenic variants that functionally confer increased cancer risk. Rather than benchmarking model performance on a surrogate metric (e.g., ClinVar variant annotations or experimental assays), we aimed to directly assesses the clinical outcome of interest (in this case, risk of breast cancer). This would also enable us to evaluate the utility of the deep learning models when applied ClinVar VUSs – a clinically relevant benchmark that by definition could not be assessed in prior studies that cross-referenced model predictions against ClinVar annotations.

Of the 254,523 female UK Biobank participants meeting inclusion criteria, 11,496 (4.52%) had been diagnosed with breast cancer at the time of analysis. As a positive control, we first constructed Firth’s penalized multivariable logistic regression models including *BRCA1* variant pathogenicity (as defined by ClinVar, comparing pathogenic vs benign variants) and age at enrollment in the UK Biobank as covariates. As anticipated, we found that pathogenic *BRCA1* variant status and older age at time of study enrollment were both associated with increased risk of breast cancer in the multivariable model (**Figure 2a; Table S3**). In these analyses, positive β regression coefficients denote increased cancer risk in pathogenic variant carriers compared to benign variant carriers. Further validating our approach, ClinVar-defined pathogenic variants in *BRCA2, ATM* and *CHEK2* were associated with increased breast cancer risk (**Figure 2a**). Additionally, ClinVar pathogenic variants in *BRCA1* and *BRCA2* conferred increased risk of ovarian/fallopian cancers (**Figure 2b**).

**Figure 2:**
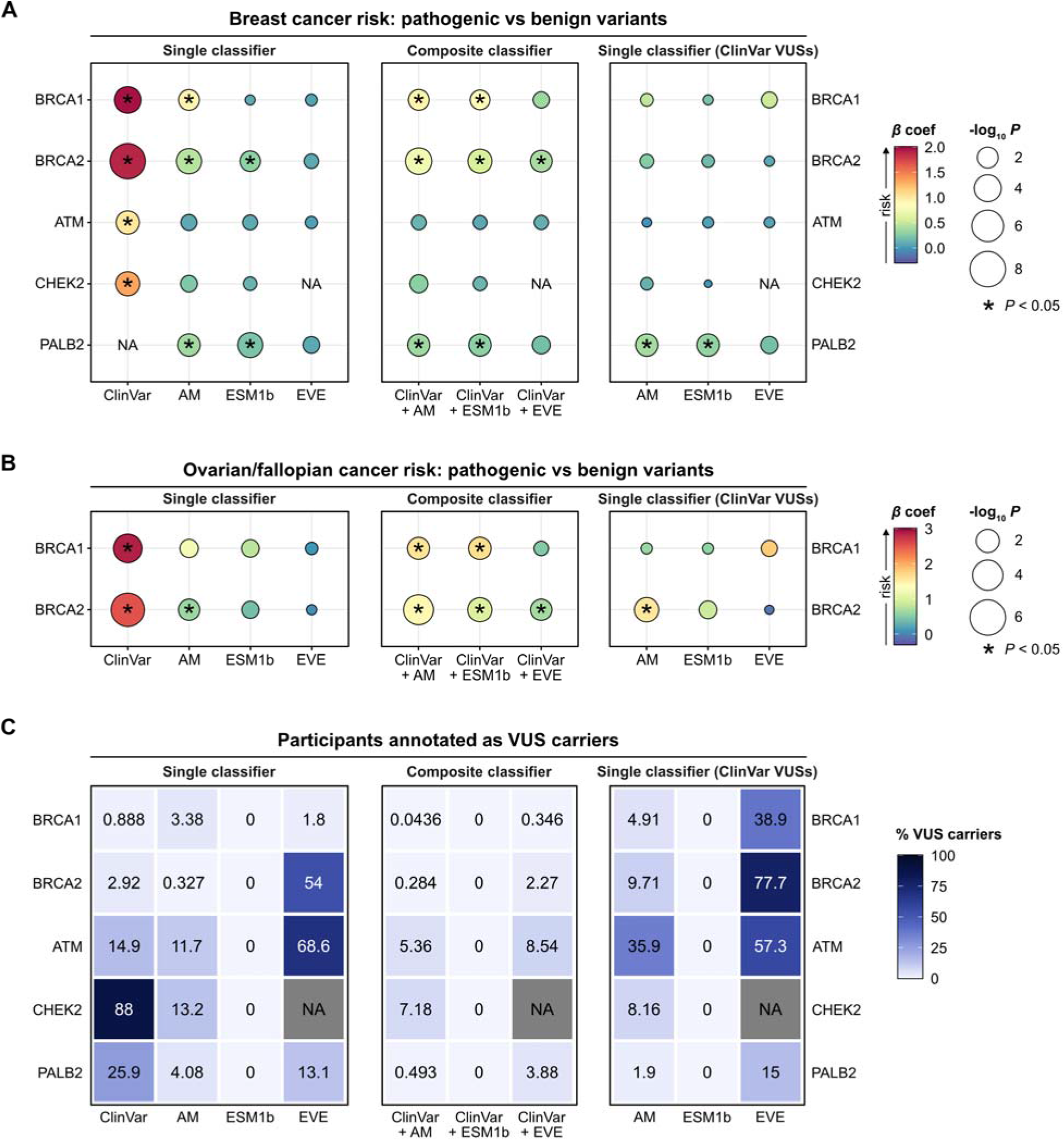
Deep learning models exhibit variable performance across genes in identifying functional pathogenic variants and are of limited utility for classifying VUSs. **A-B**. Summary statistics from Firth’s penalized logistic regression models, evaluating the association between risk of breast cancer (**A**) or ovarian/fallopian cancer (**B**) with variant pathogenicity, as determined by ClinVar or each deep learning classifier (left column). Composite classifiers were created by augmenting ClinVar annotations with model-based predictions for VUSs (middle column). Each deep learning model was also specifically evaluated on participants carrying ClinVar VUSs (right column). Points are color-coded by the magnitude of the *β* regression coefficient, with positive values indicating increased cancer risk. Points are also size-scaled by statistical significance, expressed as -log_10_ *p*-values. All models included participant age at the time of UK Biobank enrollment as a covariate. **C**. The percentage of participants annotated as VUS carriers when applying each of the different classifiers. Variants annotated as “ambiguous” by the deep learning models are counted as VUSs. Data in the right-hand panel indicates the percentage of participants classified as ClinVar VUS carriers that are then further classified by the deep learning model as carriers of an ambiguous variant.

We then utilized the pathogenicity predictions from each of the deep learning models to assess their performance in discriminating pathogenic variants that functionally confer increased breast cancer risk. AlphaMissense (but not ESM1b or EVE) was able to distinguish pathogenic *BRCA1* variant carriers with higher risk of breast cancer (**Figure 2a**), but not ovarian/fallopian cancer (**Figure 2b**). For *BRCA2*, both AlphaMissense and ESM1b identified pathogenic variants that conferred increased risk of breast cancer (**Figure 2a**), while AlphaMissense alone identified pathogenic variants associated with ovarian/fallopian cancer risk (**Figure 2b**). Of note, the effect sizes with model-based variant classifications were decreased compared to ClinVar annotations alone.

Whereas current ClinVar annotations were able to distinguish *ATM* and *CHEK2* pathogenic variants associated with increased breast cancer risk, all three deep learning models were of limited utility in this setting (**Figure 2a**). On the other hand, as none of the *PALB2* missense variants were concordantly annotated as pathogenic by ClinVar, the association of ClinVar pathogenicity labels with breast cancer risk could not be assessed. Notably, AlphaMissense and ESM1b were both able to identify *PALB2* pathogenic variant carriers with increased risk of breast cancer (**Figure 2a**), demonstrating a concrete scenario in which the deep learning models could be used to enhance clinical decision-making.

To more closely simulate how deep learning models might eventually be applied in clinical practice, we next constructed composite classifiers by augmenting existing ClinVar classifications with model-based pathogenicity predictions specifically for ClinVar VUSs. The resulting composite classifiers using either AlphaMissense or ESM1b were able to distinguish pathogenic variants in *BRCA1, BRCA2*, and *PALB2* that were associated with higher risk of breast and/or ovarian/fallopian cancer (**Figure 2a-b**), while also reducing the proportion of participants annotated as VUS carriers **(Figure 2c**). For instance, 0.888% of *BRCA1* missense variant carriers were originally annotated by ClinVar as VUS carriers, which decreased to 0.0436% when using the AlphaMissense composite classifier and to 0% with the ESM1b classifier (ESM1b utilizes a single threshold to define pathogenic vs benign variants, such that no variants are annotated as VUSs).

While the above analyses demonstrated that the deep learning models could effectively identify pathogenic variants that confer increased cancer risk while further reducing the proportion of participants annotated as VUS carriers, we observed that the composite classifiers generally had reduced discriminative power compared to ClinVar annotations alone, suggesting that the model-based classifications had diluted the specificity of ClinVar annotations. This was especially apparent in the case of *ATM* and *CHEK2*, as the ClinVar annotations in isolation were able to distinguish functionally pathogenic variants, whereas the composite classifiers could not **(Figure 2a**). Thus, we next evaluated the deep learning models exclusively on ClinVar VUS carriers. In this setting, we found that none of the deep learning models could effectively distinguish pathogenic variants in *BRCA1, BRCA2, ATM*, or *CHEK2* that conferred increased breast cancer risk (**Figure 2a**). We found similar results when assessing ovarian/fallopian cancer risk, apart from AlphaMissense when applied to *BRCA2* VUSs (**Figure 2b**). Remarkably, however, when we specifically examined *PALB2* VUS carriers (as defined by ClinVar), AlphaMissense and ESM1b both maintained discriminative power for distinguishing *PALB2* variants associated with increased breast cancer risk.

In light of the variation in model performance across genes, we wondered whether gene-specific thresholds might be helpful in improving discriminative power to identify pathogenic variants associated with cancer risk. Of note, all three of the tested deep learning models generate continuous pathogenicity scores that are subsequently partitioned into pathogenicity predictions using a common threshold across all genes. In an exploratory analysis, we tested a range of AlphaMissense score thresholds to ascertain whether more stringent cutoffs might improve model performance. For *ATM* and *CHEK2*, while the default score threshold had failed to enrich for variants associated with increased breast cancer risk (**Figure 2a**), we observed that higher thresholds could achieve statistical significance with improved effect sizes (**Figure 3a-b**). In the case of *PALB2*, higher AlphaMissense score thresholds further improved discriminative power (**Figure 3c**). We found similar results with *BRCA1* and *BRCA2*, observing that AlphaMissense classifications using more stringent score thresholds could achieve discriminative power comparable to ClinVar annotations (**Figure 3d-g**).

**Figure 3:**
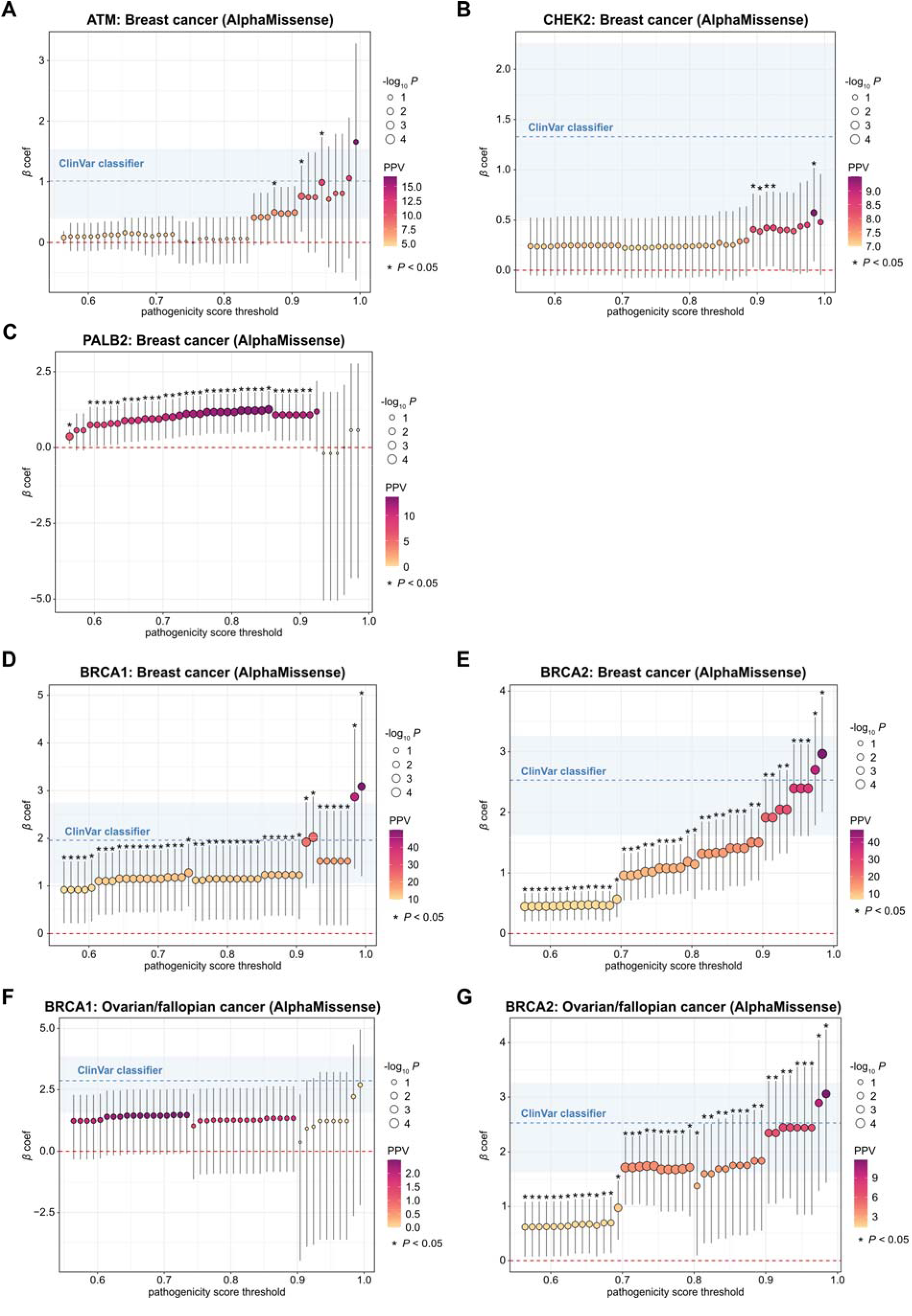
Gene-specific thresholds for defining variant pathogenicity can improve model performance. **A-G**. Summary statistics from Firth’s penalized logistic regression models with 95% confidence intervals (CIs) shown, evaluating the association between risk of breast cancer (**A-E**) or ovarian/fallopian cancer **(F-G**) with variant pathogenicity, defined by applying a range of AlphaMissense score thresholds to *ATM* (**A**), *CHEK2* (**B**), *PALB2* (**C**), *BRCA1* (**D, F**), and *BRCA2* (**E, G**). For comparison, the regression model using ClinVar variant annotations is in blue, with shaded 95% CIs. Points are color-coded by the positive predictive value (PPV) for carrying a predicted pathogenic variant. Points are also size-scaled by statistical significance, expressed as -log_10_ *p*-values. All models included participant age at the time of UK Biobank enrollment as a covariate.

In summary, here we investigated the utility of three state-of-the-art deep learning models – AlphaMissense, EVE, and ESM1b – for classifying missense variants in hereditary breast cancer genes, mimicking the clinical scenario of adjudicating cancer risk based on the results of germline genetic profiling. Overall, while we found that current deep learning models for pathogenicity prediction have potential for clinical use, their performance varied widely across genes. As a step towards addressing this problem, we explored gene-specific score thresholds and found that more stringent score cutoffs for defining pathogenic variants could potentially enhance model performance.

As presently implemented, the pathogenicity prediction models have limited utility in performing the critical task that they were designed to address – namely, the interpretation of VUSs. There is one notable exception, however: out of the five genes that we studied here, *PALB2* was unique in that none of the identified missense variants were concordantly annotated as pathogenic by ClinVar. By resolving *PALB2* VUSs into benign vs pathogenic categories, AlphaMissense and ESM1b were both able to identify *PALB2* variants functionally associated with increased risk of breast cancer. The success in classifying *PALB2* variants thus represents a key proof-of-principle for how deep learning models could eventually be leveraged to fill existing knowledge gaps and thus enhance clinical decision-making.

More generally, our findings demonstrate the value of using large-scale clinicogenomic cohorts such as the UK Biobank as a testing ground to evaluate the real-world clinical utility of pathogenicity prediction algorithms. While we focused on hereditary breast cancer genes in the present study, we expect that this approach could be readily generalized to evaluate pathogenicity predictions for genetically-mediated disorders extending far beyond cancer. Future studies on pathogenicity classification should directly benchmark their predictions on clinical outcomes such as disease incidence, as these are ultimately the endpoints that will matter to patients and clinicians.

## Methods

### Study Design and Participants

We analyzed data from the UK Biobank, a prospective cohort study comprising approximately 500,000 participants aged 37-73 years old living in the UK that were recruited between 2006 and 2010. For this study, all participants with matched exome sequencing profiles and linked health record data were included (total n = 469,623 participants). All participants provided written informed consent, which was approved by the North West Multicenter Research Ethics Committee. As the present study involved reanalysis of fully de-identified preexisting data, no additional approval was required.

### Defining cancer diagnoses

Participants in the UK Biobank were linked to national cancer registries, included as data fields 40006 (ICD10) and 40013 (ICD9). Using the UK Biobank Research Analysis Platform (RAP), we queried the entire cohort for all instances of the “Type of cancer” entry within the cancer register data, annotated by ICD9 and ICD10 codes. Breast cancer diagnoses were defined by ICD9 (174*) and/or ICD10 (C50.* or D48.6). For *BRCA1* and *BRCA2* analyses, we also considered diagnoses of ovarian or fallopian tube cancers: ICD9 (1830, 1832) and/or ICD10 (C56, C57.0, C57.4, D39.1). All participants with matched genomic and clinical data were included for the analyses comparing variant pathogenicity annotations between ClinVar and each deep learning model. For analyses of breast or ovarian cancer risk in relation to pathogenic vs benign variant carrier status, only female participants were selected.

### Analysis of exome sequencing data

Within the UK Biobank RAP, we used the Swiss Army Knife tool to filter and annotate variants in *BRCA1, BRCA2, ATM, CHEK2*, and *PALB2*. Specifically, we used PLINK2 to extract variants in the chromosome regions corresponding to each of the 5 target genes, with a minimum minor allele frequency of 0, a minimum minor allele count of 4, a Hardy-Weinberg equilibrium filter of *p* < 1*10^−15^ with the “keep-fewhet” flag activated, as well as less than 10% of missing genotypes and variant calls.^13^ From there, we used SNPEFF^14^ to annotate each variant by their functional type and impact on protein sequence, with further annotation by SNPSIFT^15^ to incorporate annotations from the February 15, 2024 ClinVar data release.

ClinVar^16^ labels were consolidated into benign, pathogenic or VUS categories, with conflicting or missing annotations consolidated as VUSs. For all identified missense coding variants, we used precomputed scores from AlphaMissense^7^, EVE^8^ and ESM1b^9^ to predict variant pathogenicity. As the EVE database (https://evemodel.org/) did not have predictions available for *CHEK2*, EVE was excluded from the *CHEK2* analyses. We examined UK Biobank participants with at least one missense variant in each of the five genes and classified the participants as benign, VUS, or pathogenic variant carriers. Given that individual participants may have multiple variants simultaneously, pathogenic variants were prioritized over VUSs, and in turn VUSs were prioritized over benign variants. For instance, if a participant had both pathogenic and benign variants in a given gene, the participant was annotated as a pathogenic variant carrier and not as a benign variant carrier.

We then compared ClinVar and the deep learning model annotations, first on the level of unique variants and subsequently on the participant level to evaluate the accuracy of the deep learning models in recapitulating ClinVar pathogenicity labels. To model a clinical scenario in which deep learning models might be applied, we generated composite classifiers by starting with ClinVar annotations as a foundation and augmenting them with model-based pathogenicity predictions to specifically classify ClinVar VUSs; in other words, all benign and pathogenic ClinVar annotations were retained for the composite classifiers, and the pathogenicity predictions were only applied to ClinVar VUSs.

### Association of variant pathogenicity with cancer risk

To assess whether the pathogenicity classifications were functionally associated with breast cancer risk, we analyzed female participants with a benign vs pathogenic missense variant for the gene of interest, while excluding participants that carried a frameshift, stop gain, stop loss, or start loss variant in that particular gene. We then used Firth’s penalized logistic regression to determine the association of pathogenic vs benign variants with diagnoses of breast or ovarian/fallopian cancer. In all regression models, we included age at time of enrollment in the UK Biobank as a covariate, binarizing by the median age. Participants carrying a VUS (as defined by ClinVar or each deep learning model), but no pathogenic variants, were excluded from the regression analysis, regardless of the presence of co-occurring benign variants (see discussion above). We further calculated regression models using the composite pathogenicity labels (see above) to simulate the clinical scenario of first relying on ClinVar annotations where available and subsequently employing deep learning models to classify VUSs. To directly assess the predictive utility of deep learning models on classifying VUSs, we analyzed VUS carriers only (as defined by ClinVar) and assessed whether the predicted pathogenicity labels were associated with cancer risk.

Regression results were reported as β coefficients and *p*-values, with a significance threshold of *p* < 0.05. We did not adjust for multiple comparisons. The 95% confidence intervals (CIs) for all regression β coefficients are detailed in the supplementary tables.

### Assessment of gene-specific thresholds for defining pathogenic variants

For evaluating whether gene-specific thresholds could improve model performance, we focused on AlphaMissense pathogenicity scores. We retained the default threshold of 0.34 to distinguish benign vs ambiguous variants, while varying the threshold to distinguish ambiguous vs pathogenic variants, ranging from 0.564 (the default) to 1, in increments of 0.01. Regression results were reported as described above.

## Code availability

Initial data pre-processing and variant annotation were performed on the UK Biobank RAP: https://ukbiobank.dnanexus.com/landing. Custom in-house analysis code has been deposited to Github: https://github.com/rdchow/UKB_pathogenicityPrediction.

## Data availability

All participant-level genomic and phenotypic data are available through the UK Biobank, through the standardized data-access protocol. Details for registering are available at https://www.ukbiobank.ac.uk/enable-your-research/register. Summary statistics generated in this study are available in the supplementary tables.

## Contributions

RDC conceived and designed the study. RDC performed the data analysis. RDC, RBP, and KLN prepared the manuscript. RBP secured funding. RBP and KLN jointly supervised the study.

## Acknowledgements

This study was supported by the NIH/NCI (5K08CA263541), awarded to RBP.

## Competing interests

RDC reports no competing interests. RBP has received grants from the National Institutes of Health, Department of Defense, Prostate Cancer Foundation, National Palliative Care Research Center, NCCN Foundation, Conquer Cancer Foundation, Humana, Emerson Collective, Schmidt Futures, Arnold Ventures, Mendel.ai, and Veterans Health Administration; personal fees and equity from GNS Healthcare, Thyme Care, and Onc.AI; personal fees from the ConcertAI, Cancer Study Group, Biofourmis, Genetic Chemistry Therapeutics, CreditSuisse, G1 Therapeutics, Humana, and Nanology; honoraria from Flatiron and Medscape; has board membership (unpaid) at the Coalition to Transform Advanced Care and American Cancer Society; and serves on a leadership consortium (unpaid) at the National Quality Forum, all outside the submitted work. KLN reports serving on a Scientific Advisory Board for Merck, unrelated to the current study.

## Notes

### Funding Statement

This study was funded by the NIH/NCI (5K08CA263541), awarded to RBP.

### Author Declarations

All UK Biobank participants provided written informed consent, which was approved by the North West Multicenter Research Ethics Committee. As the present study involved reanalysis of fully de-identified preexisting data, no additional approval was required.

## References

1. Couch, F. J., Nathanson, K. L. & Offit, K. Two Decades After BRCA: Setting Paradigms in Personalized Cancer Care and Prevention. Science 343, 1466–1470 (2014).

2. Domchek, S. M. et al. Association of risk-reducing surgery in BRCA1 or BRCA2 mutation carriers with cancer risk and mortality. JAMA 304, 967–975 (2010).

3. US Preventive Services Task Force. Risk Assessment, Genetic Counseling, and Genetic Testing for BRCA-Related Cancer: US Preventive Services Task Force Recommendation Statement. JAMA 322, 652–665 (2019).

4. Lindor, N. M., Goldgar, D. E., Tavtigian, S. V., Plon, S. E. & Couch, F. J. BRCA1/2 Sequence Variants of Uncertain Significance: A Primer for Providers to Assist in Discussions and in Medical Management. Oncologist 18, 518–524 (2013).

5. Makhnoon, S., Bednar, E. M., Krause, K. J., Peterson, S. K. & Lopez-Olivo, M. A. Clinical management among individuals with variant of uncertain significance in hereditary cancer: A systematic review and meta-analysis. Clinical Genetics 100, 119–131 (2021).

6. Daly, M. B. et al. Genetic/Familial High-Risk Assessment: Breast, Ovarian, and Pancreatic, Version 2.2021, NCCN Clinical Practice Guidelines in Oncology. J Natl Compr Canc Netw 19, 77–102 (2021).

7. Cheng, J. et al. Accurate proteome-wide missense variant effect prediction with AlphaMissense. Science 381, eadg7492 (2023).

8. Frazer, J. et al. Disease variant prediction with deep generative models of evolutionary data. Nature 599, 91–95 (2021).

9. Brandes, N., Goldman, G., Wang, C. H., Ye, C. J. & Ntranos, V. Genome-wide prediction of disease variant effects with a deep protein language model. Nat Genet 55, 1512–1522 (2023).

10. Bycroft, C. et al. The UK Biobank resource with deep phenotyping and genomic data. Nature 562, 203–209 (2018).

11. Sudlow, C. et al. UK biobank: an open access resource for identifying the causes of a wide range of complex diseases of middle and old age. PLoS Med 12, e1001779 (2015).

12. Breast Cancer Association Consortium et al. Breast Cancer Risk Genes - Association Analysis in More than 113,000 Women. N Engl J Med 384, 428–439 (2021).

13. Szustakowski, J. D. et al. Advancing human genetics research and drug discovery through exome sequencing of the UK Biobank. Nat Genet 53, 942–948 (2021).

14. Cingolani, P. et al. A program for annotating and predicting the effects of single nucleotide polymorphisms, SnpEff. Fly (Austin) 6, 80–92 (2012).

15. Cingolani, P. et al. Using Drosophila melanogaster as a Model for Genotoxic Chemical Mutational Studies with a New Program, SnpSift. Front Genet 3, 35 (2012).

16. Landrum, M. J. et al. ClinVar: public archive of relationships among sequence variation and human phenotype. Nucleic Acids Res 42, D980–985 (2014).

